# Hydroxychloroquine Prophylaxis against Coronavirus Disease-19: Practice Outcomes among Health-Care Workers

**DOI:** 10.1101/2021.08.02.21260750

**Authors:** Parloop Bhatt, Vishva Patel, Prachi Shah, Keyur Parikh

## Abstract

**Background:** Severe acute respiratory syndrome coronavirus 2 (SARS-CoV-2) is a rapidly emerging virus responsible for the ongoing Covid-19 pandemic with no known effective prophylaxis. We investigated whether hydroxychloroquine(HCQ) could prevent SARS CoV-2 in healthcare workers(HCW) at high-risk of exposure.

**Method:** This voluntary observational study for the prevention and treatment of COVID-19 was conducted at a tertiary care center, from 12^th^ June to 12th October 2020(total 16 weeks). All consented asymptomatic HCW’s of CIMS hospital were administered 400 mg HCQ twice a day on day one followed by 400 mg once weekly to be taken with meals up to 16 weeks. Data collected included OPD registration, risk assessment, medical and family history (related to COVID), physical examination and vitals, pulse oximetry, ECG (pre and post HCQ), drug adherence, side effects, adverse drug reactions.

**Result:** The study enrolled 927 full-time, hospital-based HCWs ((including doctors, nurses, paramedical, lab technicians, sanitary workers and others), of whom 731(78.85%) initially started HCQ while 196 (21.14%) did not volunteer. The median age and weight of the study population was 27.5 years and 69.5 kg respectively. No major associated co-morbidities were present in these HCW’s. There was an increased trend towards non adherence to HCQ with each proceeding week more so after week 11. Of the 731 HCW’s taking HCQ a total of 167(22.8%) tested COVID positive at different intervals of time as against 30 HCW (15.3%) out of 196 not taking HCQ. The rate of COVID-19 positive was statistically significantly higher in the HCW’s taking HCQ (p=0.0220; 95% CI: 1.14% to 12.94%), as compared to those not on HCQ. Thus HCQ was not prophylactically effective against COVID 19 infection. No participants in this study experienced grade 3 or 4 adverse events. No significant difference in the median of ECG changes in QTc between pre and post HCQ administration of 46 HCW’s was observed.

**Conclusions:** This clinical study did not detect a reduction in SARS CoV-2 transmission with prophylactic administration of 400 mg/HCQ in HCW’s. All participants who did contract SARSCoV-2 were either asymptomatic or had mild disease courses with full recoveries. All adverse events were self-limiting and no serious cardiovascular events were reported with use of HCQ. In the absence of robust data, it seems premature to recommend HCQ as a prophylactic panacea for COVID-19.

## INTRODUCTION

Coronavirus disease 2019 (COVID 19) is a rapidly emerging infectious disease caused by the severe acute respiratory syndrome coronavirus 2 (SARS-CoV-2). Among contacts of persons with Covid-19, the percentage in which new cases developed (secondary attack rate) has been estimated at 10 to 15%.^[1–4]^ The etiologic agent, SARS-CoV-2 belongs to the subgenus Sarbecovirus of the genus Beta coronavirus, family Coronaviridae. Viruses of the family Coronaviridae possess a single-strand, positive-sense RNA genome. ^[5]^

The infection-control strategy before vaccines was based on non-pharmacologic interventions, including hand hygiene, use of face masks, social distancing, and isolation of case patients and contacts. ^[6,7]^ The virus spreads among close contacts mainly through respiratory droplets through contact with mucous membranes of the mouth, nose, and possibly eyes. ^[8]^ Clinical trials are underway in various parts of the globe to evaluate the efficacy of hydroxychlororquine (HCQ) chemoprophylaxis in COVID-19 among health care workers (HCW) ^[9–10]^. However, HCQ could have serious adverse effects of which the HCW should be aware. The National Task Force (NTF) for COVID 19 constituted by Indian Council of Medical Research (ICMR) under the Ministry of Health and Family Welfare (MoHFW) issued guidelines dated 21st March 2020 on prophylactic use of HCQ/Chloroquine (CQ) against SARS-CoV-2 for high risk population, including asymptomatic HCWs involved in direct care of suspected or confirmed cases of COVID 19. ^[11]^ As per revised advisory, the Joint Monitoring Group and NTF had recommended the prophylactic use of HCQ in the following categories: 1) All asymptomatic healthcare workers involved in containment and treatment of COVID19 and asymptomatic healthcare workers working in non-COVID hospitals/non-COVID areas of COVID hospitals/blocks, 2) Asymptomatic frontline workers, such as surveillance workers deployed in containment zones and paramilitary/police personnel involved in COVID-19 related activities. 3) Asymptomatic household contacts of laboratory confirmed cases. ^[12]^

Initially used to treat malaria, HCQ is an important therapeutic option for several autoimmune diseases, especially systemic lupus erythematous (SLE) and rheumatoid arthritis (RA). ^[13, 14, 15, 16]^ The efficacy of HCQ in rheumatic illnesses stems from its anti-inflammatory and immunomodulatory effects, the mechanisms of which are unclear. Although initially thought to exert its immunomodulatory effects by interfering with lysosomal enzymatic actions and major histocompatibility complex class-II (MHC-II)-mediated antigen presentation, emerging evidence suggests interference with Toll-like receptor (TLR) functions as an additional pathway. ^[17]^ Although overall efficacy of HCQ in infectious diseases, besides malaria, is unknown, HCQ is being explored in human immunodeficiency viruses, Coxiella burnetii, Zika virus, chikungunya, and Whipple’s disease. ^[15]^ The mechanism of action by which HCQ exhibit antiviral properties against SARS-CoV-2 has not been fully elucidated but it is presumed that it is a weak base that concentrates on the intracellular sections including endosome and lysosome; so, viral replication in the phase of fusion and un-coating is stopped. Also, it can change the ACE2 glycosylation and inhibit both S protein binding and phagocytosis. The last mechanism would be the suppressing effect on cytokine production and the immunomodulatory effect of the drug. ^[18, 19]^ In vitro effect of HCQ on SARS-CoV-2 infected Vero-cells using physiologically based pharmacokinetic models showed inhibitory effect of HCQ on entry and post-entry steps of viral replication. ^[20,21]^ However, rare but serious adverse effects have been reported, mostly with long term use. HCQ-induced acquired lysosomal storage disease causes some of these adverse effects, including myopathy and cardiomyopathy. ^[1]^ Corrected QT (QTc) interval prolongation is associated with HCQ owing to human ether-a-go-go-related gene (hERG) voltage-gated potassium channel inhibition. ^[22]^

HCQ sulfate (blood half-life 537 hours or 22.4 days) attains peak blood levels 3.26 hours after administration of 200 mg salt (155-mg base) orally in healthy males. ^[23]^ Absorption of the drug was found to be less in patients with RA with severe disease activity compared with the less severe groups. This observation may have significant importance while ascertaining dosage recommendation in healthy subset of population. ^[24]^

As per various studies reported on role of HCQ as a prophylaxis in COVID-19, there is lack of robust studies to support the use of HCQ as a chemo prophylactic agent for pre-exposure prophylaxis. As well there is also a concern regarding the adverse effects associated with it. Some studies suggested the use of HCQ as a prophylaxis as it has certain good characteristics like favorable pharmacokinetics, low cost, readily available, in vitro evidence of benefit. While some of the studies are not in favor of use of HCQ as prophylaxis in COVID-19 as currently used dosing regimens may be inadequate to provide a benefit, although higher doses may lead to significant safety concerns.

In context to the above literature, a study was designed to evaluate the safety and efficacy of HCQ as prophylactic treatment for HCW deployed in non-COVID as well as COVID areas at a COVID care hospital.

## METHOD

This is an observational study to establish the effectiveness of a weekly prophylactic regime of HCQ in prevention of COVID-19 in HCW. The study was conducted at CIMS hospital, Ahmedabad, India which is an Ahmedabad Municipal Corporation approved COVID care hospital. The protocol was approved by the institutional ethics committee (#ECR/206/Inst/GJ/2013/RR-20). HCW’s were categorized as “Clinical HCW” defined as those HCWs involved in direct patient care (COVID or non COVID) as part of their regular routine, and “Nonclinical HCW” defined as those not involved in patient care and with no opportunity for patient contact during their regular work routine.

All HCW irrespective of their HCQ consumption status, who were asymptomatic at baseline and were willing to give consent and take HCQ as recommended were included in the study. The study period extended from 12^th^ June to 12th October 2020, total 16 weeks (including screening visit or pretreatment visit). HCQ was administered only on the prescription of a registered medical practitioner.

All consented asymptomatic healthcare workers of CIMS hospital were administered 400 mg HCQ twice a day on day one followed by 400 mg once weekly to be taken with meals up to 16 weeks.

Data collected included OPD registration, risk assessment, medical and family history (related to COVID), physical examination, blood pressure, heart rate, pulse oximetry, respiratory rate, ECG, drug adherence, side effects, adverse drug reactions (ADR). ECG was performed prior to study medication (HCQ) and at 4 weeks post treatment. The primary outcome was the incidence of SARS-CoV-2 infection as determined by RT-PCR through nasopharyngeal swab during HCQ treatment. Secondary outcomes included adverse effects, treatment discontinuation, frequency of QTc prolongation, and clinical outcomes for SARS-CoV-2–positive participants.

### Ethical Consideration

The study was approved by the CIMS (Care Institute of Medical Sciences) hospital ethics committee (#OC-57/5th June 2020).

### Statistical Plan

Demographics and baseline characteristics (age, gender etc.), ECG – QT_c_ changes were summarized using descriptive statistics. Categorical variables were summarized using frequencies and percentages. All analyses were performed using STATA, version 11.0. Continuous variables were reported as medians. Means and standard deviation (±SD) were determined for frequency variables. For statistical significance, measures of effect; odds ratios with 95% CI’s, and a 2-sided p value less than 0.05 were applied. Two proportion values were analyzed with Chi-square test and mean with standard deviation was analyzed with two samples t-test.

## RESULTS

Of the **t**otal 927 HCW at CIMS hospital 731 (78.9%) HCW (median age 27.5 years, range: 20-52 years) voluntarily consented for participation in the study and were administered HCQ. Of these 731 HCW, 552 were clinical HCW while 179 belonged to non-clinical category. Table 1 shows demographic and associated risk factor details. Median weight of these HCW was 69.5 kg weight. Majority had no associated co-morbidity.

**Table: 1.** Demographic and associated risk factors among HCW.

| **Characteristic** | **All HCWs**<br>**(N=927)**<br>**N (%)** | **HCWs on HCQ**<br>**N=731 (78.86%)**<br>**N (%)** | **HCWs not on HCQ**<br>**N=196 (21.14%)**<br>**N (%)** |
| --- | --- | --- | --- |
| **Age, median(range),y** | 27.5 | 28 | 27 |
| **Weight, median(range),kg** | 69.5 | 69 | 70 |
| **BMI, median(range)** | 26.5 | 26 | 27 |
| **Women** | 363 (39.16) | 271(37.07) | 92(46.94) |
| **Current smoker** | 7(0.76) | 7(0.96) | 0 |
| **Asthma** | 3(0.32) | 3(0.41) | 0 |
| **Diabetes** | 6(0.65) | 5(0.68) | 0 |
| **Hypertension** | 9(0.97) | 8(1.09) | 1(0.51) |
| **Thyroid** | 5(0.54) | 5(0.68) | 0 |
| **Allergic** | 3(0.32) | 2(0.27) | 0 |
| **None** | 901(97.20) | 706(96.58) | 195(99.49) |
| **Family HT** | 37(3.99) | 33(4.5) | 4(2.04) |
| **Family Diabetes** | 28 | 25(3.41) | 3(1.5) |
| **Family heart Disease** | 19 | 17(2.32) | 2(1.02) |
| **Family TB** | 3 | 2(0.27) | 1(0.51) |

### HCQ Adherence and Prophylactic Efficacy of HCQ against COVID19

Figure 1 describes weekly adherence and efficacy to prophylactic use of HCQ by HCW’s over a period of 16 weeks. Of the 937 HCW’s, 731 HCW’s voluntarily started HCQ, however 166 discontinued at week1.At week 2 of the 565 HCW’s, 46 discontinued HCQ. At 7^th^ week, 295 out of 426 discontinued HCQ. By 11^th^ week nearly all discontinued HCQ except 5 which did not continue beyond 16^th^ week. There was an increased trend towards non adherence to HCQ with each proceeding week more so after week 11. Fig.1 also depicts HCW’s who reported COVID positive at respective weeks after taking HCQ. No specific trend related to time was observed for HCWs reporting COVID positive at each week. However, number of HCW’s testing COVID positive was higher in those working clinically more so the clinical care staff nurses (60%) as compared to non-clinical staff irrespective of HCQ use. All COVID positive HCW’s showed typical signs of infection like fever, fatigue, headache, sore throat, shortness of breath etc. whose severity was unaffected by use of HCQ.

**Fig. 1:**
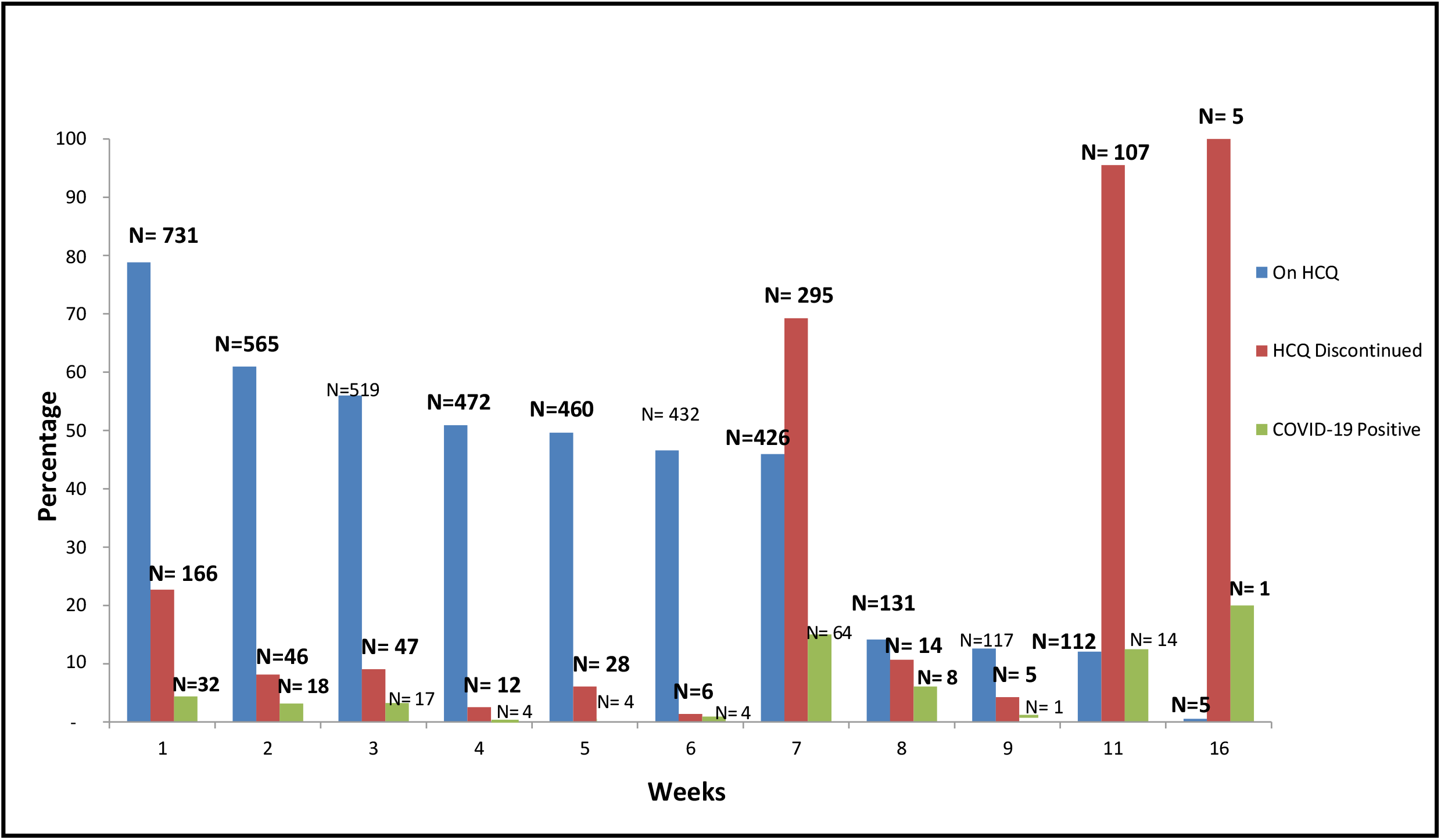
Weekly Status of HCQ Adherence and Positive COVID-19 infection in HCW’s.

### Efficacy of HCQ against COVID-19 on HCW

Of the 731 HCW’s taking HCQ a total of 167(22.8%) tested COVID positive at different intervals of time as against 30 HCW (15.3%) out of 196 not taking HCQ (Fig.2) The rate of COVID-19 positive was statistically significantly higher in the HCW’s taking HCQ (22.84% vs. 15.30% p=0.0220; 95% CI: 1.14% to 12.94%), as compared to those not on HCQ. Thus HCQ was not prophylactically effective against COVID 19 infection.

**Fig 2:**
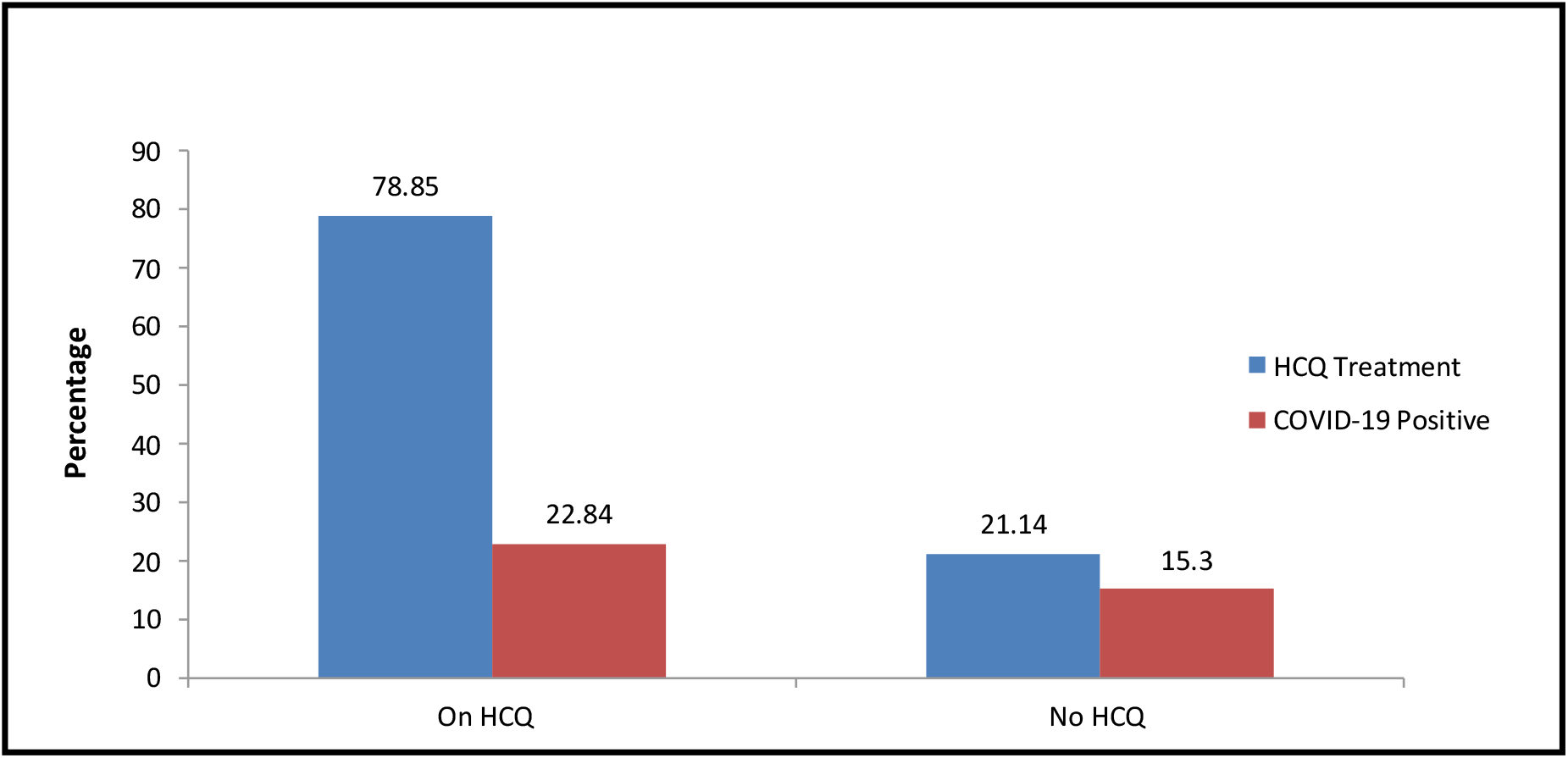
Efficacy of HCQ against COVID-19 on HCW.

### Side Effects and Adverse Events of HCQ in HCW

No participant in this study experienced grade 3 or 4 adverse events on the Common Toxicity Criteria for Adverse Events scale. The safety profile analysis in terms of side effects and adverse event noted were 259 of participating HCW experiencing at least one side effect/ adverse event following use of HCQ. About 90.4 % HCW’s experienced no adverse event. All effects and events were mild to moderate in nature, requiring no hospitalization. Most common side effects observed were headache (9.8%), followed by nausea and vomiting (6.9%), GI distress (3.4%) and weakness (2.7%). About 1.5 % experienced fever or chest pain or body pain. No cardiac events (e.g. syncope and arrhythmias) were observed.

ECG was performed for 46 HCWs pre and post use of HCQ i.e. at week 0 and week 4. There was no significant difference in the median of changes in QTc between pre and post HCQ administration (4.2 milliseconds vs 4.3 milliseconds; 95% CI, −4.66 to 9.22; Chi-square 2-sample *t* test, *P* =0.51).

## DISCUSSION

Medication-based prevention and treatment of COVID-19 is a challenge worldwide. No pharmacologic prophylaxis for COVID-19 has been established. Given the high exposure of HCWs to COVID-19 worldwide, there was great interest in determining the prophylactic effectiveness of a medication to prevent viral transmission. Zhou et al. (2020) published results supporting the prophylactic use of HCQ because of its *in vitro* ability to interfere with cellular receptors for COVID-19 and block virus fusion with host cells. ^[25]^ During the COVID-19 emergency, HCQ has been prescribed as off-label treatment, with several differences between countries. Many regulatory authorities have issued warnings on HCQ safety based on some retrospective studies. ^[26]^ The advantage of using HCQ is that its toxicity profile is well known thus fewer commercial, legal and financial constraints. ^**[27]**^ However robust clinical evidence is still awaited.

Since the study site was a COVID care hospital, a pre-exposure prophylaxis strategy was followed where it was considered that prevention possibly depended on dose and duration of therapy, since determining cause of true exposure in transmission was challenging.

A loading dose of 400–800 mg was used in 84% of the clinical trials placed on clinicaltrials.gov.^.[28]^ Indicating uncertainty and conflicts clinical trial results depict heterogeneous doses ranging from 200 to 600 mg/day (50.6%) and from 200 to 400 mg/week (24.7.%).^[29]^At our study site following ICMR guidelines HCQ was administered 400mg twice on day 1 and 400 mg every week.^[11,12].^Since heterogeneous and unjustifiable *in vitro* duration data is available we continued gathering data from HCW’s up to 16 weeks of HCQ treatment. Although the number of HCW’s non-adherence to HCQ steadfastly increased. Of the 927 HCWs associated with the hospital, 731 voluntarily consented to participate. All were well informed about the safety and efficacy of HCQ. The baseline characteristics of HCWs revealed all were healthy with no comorbidities. Every Wednesday was considered as a HCQ dose day during study period.

With every proceeding week adherence towards HCQ declined. The reason of non-adherence was not ascertained however efforts like weekly online sessions from human resource department were placed emphasizing about HCQ adherence. By week 7, nearly 50% HCW’s had stopped HCQ. HCQ has well documented adverse effect profiles including but not limited to gastrointestinal upset and headache.^[30]^ More serious adverse effects such as QTc interval prolongation on ECG, cardiac arrhythmias, and retinopathy ^[31]^ are associated with chronic therapy.^[32]^ However, in general, HCQ is considered to be safe, and side effects are generally mild and transitory, despite the margin between the therapeutic and toxic dose being narrow. No participants in this study experienced grade 3 or 4 adverse events on the Common Toxicity Criteria for Adverse Events scale, hospitalizations, or death. About 32 % of our study participants experienced mild side effects subsequent to the prophylaxis, but the drug was more or less well tolerated. In those who experienced side effects, gastrointestinal symptoms such as gastric irritation, nausea, vomiting were the most common ones. Few also reported headaches and dizziness. With regard to safety issues, our results are similar to Abella et al. wherein no grade 3 or four adverse events or cardiac events were reported unlike the study of Rajasingham et al. ^[33,34]^ which reported one cardiac event that was potentially HCQ-related. Although all HCWs who took HCQ/CQ prophylaxis were aware of the side effects but still very few of them did baseline investigation like ECG, to rule out prolonged QT, which is the common adverse effect of the drug. Data of 46 HCW’s pre and post HCQ prophylaxis reported no significant difference in the median of changes in QTc (P =0.51).

In our study no reduction in SARS CoV-2 infection with prophylactic use of HCQ was noted. Of the 731 HCWs on HCQ, 167(22.8%) tested COVID positive at different intervals of time as against 30 HCW (15.3%) out of 196 not taking HCQ. The rate of COVID 19 in HCWs taking HCQ was not less as compared to those not taking HCQ with infections occurring throughout the 16-week period. The conversion of HCWs to SARS-CoV-2 positive status was determined either if the HCW developed symptoms and was referred for a nasopharyngeal swab test or his close contact tested positive and was thus referred for nasopharyngeal test and tested positive at any point of time in 16 weeks. All COVID positive HCWs irrespective of use of HCQ were either asymptomatic or had mild disease and fully recovered. Our results fall in line with the observations conducted by Rajasingham who reported no significant reduction in COVID incidence in any of the three arms ((HCQ 400 mg weekly *vs*. HCQ 400 mg twice weekly vs. placebo). ^[34]^ Also Abella et al. carried out a randomized double-blind placebo-controlled trial on a HCW population, reporting no significant differences in infection rates between participants taking HCQ or placebo. The trial was stopped early for futility before reaching the planned enrollment. ^[33]^

## Conclusion

This clinical study did not detect a reduction in SARS CoV-2 transmission with prophylactic administration of HCQ in HCWs. All participants who did contract SARSCoV-2 were either asymptomatic or had mild disease courses with full recoveries. All adverse events were self-limiting and no serious cardiovascular events were reported with use of HCQ. In the absence of a robust data, it seems premature to recommend HCQ as a prophylactic panacea for COVID-19.

## Data Availability

The authors and institute affirms that the manuscript is an honest, accurate, and transparent account of the data being reported. No available important aspects of the study have been omitted.

